# The Potential Impact of Interruptions to HIV Services: A Modelling Case Study for South Africa

**DOI:** 10.1101/2020.04.22.20075861

**Authors:** Britta Jewell, Jennifer Smith, Timothy Hallett

## Abstract

The numbers of deaths caused by HIV could increase substantially if the COVID-19 epidemic leads to interruptions in the availability of HIV services. We compare publicly available scenarios for COVID-19 mortality with predicted additional HIV-related mortality based on assumptions about possible interruptions in HIV programs. An interruption in the supply of ART for 40% of those on ART for 3 months could cause a number of deaths on the same order of magnitude as the number that are anticipated to be saved from COVID-19 through social distancing measures. In contrast, if the disruption can be managed such that the supply and usage of ART is maintained, the increase in AIDS deaths would be limited to 1% over five years, although this could still be accompanied by substantial increases in new HIV infections if there are reductions in VMMC, oral PrEP use, and condom availability.

## Background

In addition to the direct health impact of the COVID-19 epidemic, there may also be a detrimental impact on service provision for other health conditions due to increasing demands on overall health service capacity, interruptions to supply of medicines, or funding shortages. Here we examine the impact of potential hypothetical disruption to HIV services in South Africa. Interruptions to different elements of the HIV treatment and prevention cascades have differential impacts on the resulting loss of health and it will be useful for decision-makers to understand the relative impact of different reductions in service in order to inform planning for service continuity during the COVID-19 epidemic.

## Methods

We hypothesised three patterns of disruption to HIV services in South Africa that might occur during the COVID-19 epidemic, described below and summarised in Table 1. Each scenario is cumulative in the sense that it incorporates the changes described in all less severe scenarios. In the first instance, we assume the disruption begins in mid-April 2020 and lasts for 3 months, and that thereafter normal service resumes for all programs.

**Table 1:**
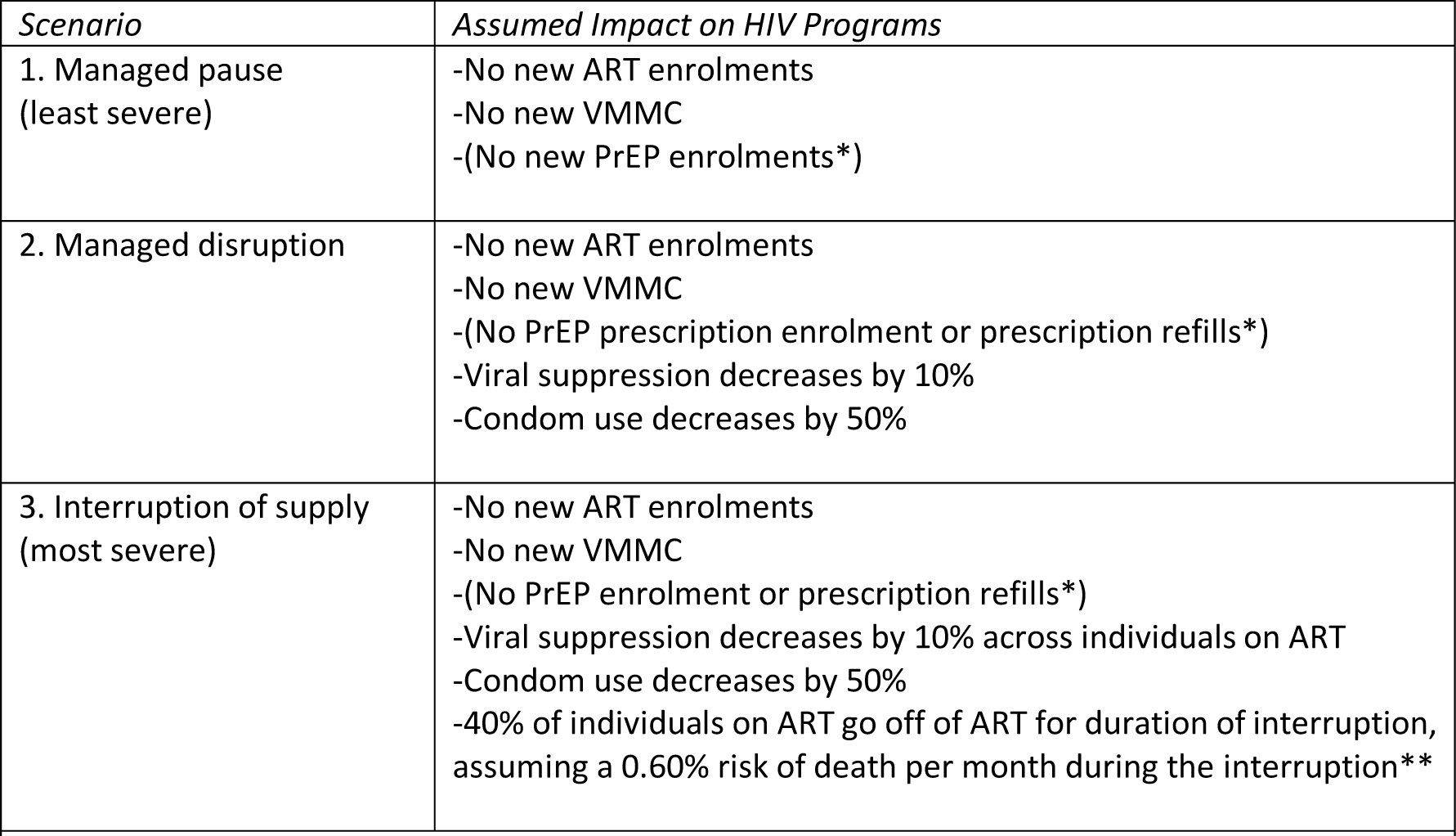
Scenarios characterising disruption to HIV services due to the COVID-19 epidemic. *PrEP coverage is assumed to be small and does not affect directly results presented here. **We optimistically assume that if individuals have stopped ART use due to the COVIDL19 disruption, they immediately return to ART once supply resumes and their long-term prognosis on ART is not affected by the interruption in ART usage.

1. *Managed pause* (least severe): In this scenario, the expansion of services planned for this time is paused but all services are maintained at their current levels. This might occur, for instance, if there is very little actual disruption to the HIV programs, but opportunities to expand are not available due to it being impractical to run outreach services and persons not yet engaged with care postponing testing.
2. *Managed disruption:* In this scenario, substantial pressure on the health system and social distancing measures combine to disrupt services, but these are sufficiently well managed that their worst impacts are mitigated, and the supply of ART is maintained. In addition to the effects of the first scenario (‘Managed pause’) there is also a reduction in condom use, as might be caused by reduced supply of condoms and less opportunity to acquire them, and decreased viral suppression, as might be caused by a combination of persons being less inclined or able to present for routine viral load testing, viral load testing not being so widely available (due to increased usage of laboratory equipment), and short-term fluctuations in the supply of ART drugs.
3. *Interruption of supply* (most severe): In this scenario, extreme pressure on the health system and/or strict interventions and stressed supply chains domestically and internationally combine to interrupt the supply of key medicines, with the result that a fraction of people living with HIV (PLHIV) on ART are forced off ART.

We used an established deterministic mathematical model of the HIV epidemic in South Africa to quantify the impact of these disruptions over the five-year period, 2020-2024.[1, 2] The number of ‘excess’ outcomes (deaths and life-years lost) that are attributed to these disruptions are computed in reference to a model projection in which no interruptions occur and the coverage of programs expands in the manner that would have otherwise been anticipated.

We compared the effect on HIV programmes to the anticipated direct effects from a COVID-19 epidemic in South Africa (Table 2).[3]

**Table 2:**
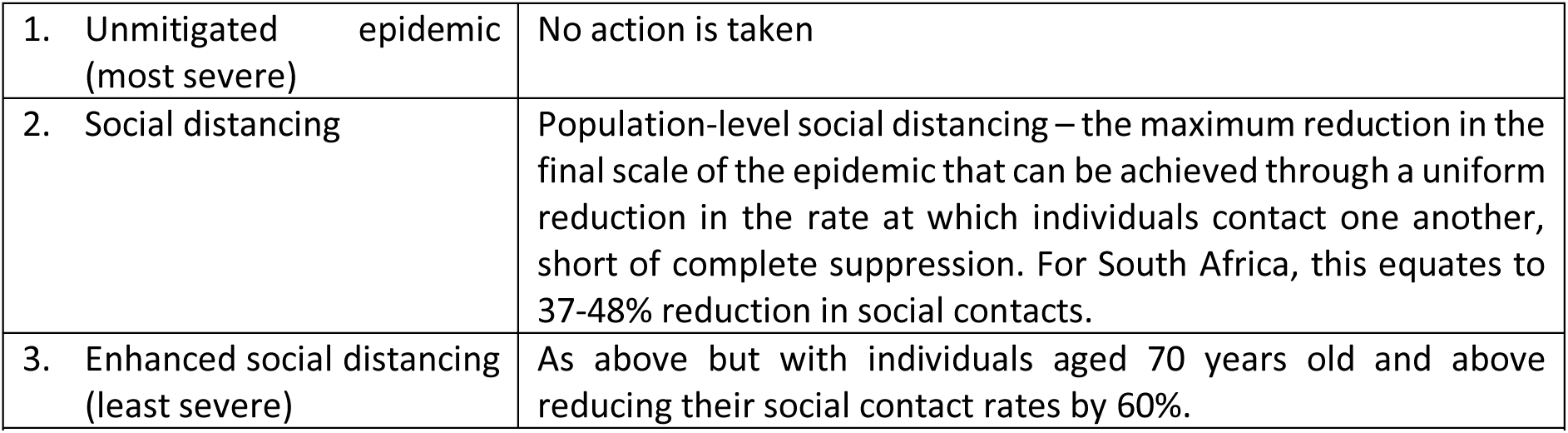
Public health responses to the COVID-19 epidemic (from [3]).

Results are most sensitive to the risk of dying for PLHIV who have had their ART supply interrupted. This quantity is not well known or understood. For the main analysis, we assume an average monthly risk of death of 0.60% for this group. We conducted a sensitivity analysis using three different values for this value (Table 3). We also note that the monthly risk of death is likely to increase over time as individuals accrue time off of ART; however, this is not represented in the model.

**Table 3:** Alternative assumptions used for the risk of death experienced by those PLHIV whose ART supply is interrupted.

|  | Monthly mortality risk | Proportion that would die after one year | Justification |
| --- | --- | --- | --- |
| Lower bound | 0.26% | 3% | The SMART trial found a 3% risk at 12 months of either death of an opportunistic infection for those with interrupted ART.[4] |
| Medium | 0.60% | 7% | This implies a mean survival of 14 years which is approximately the survival time for HIV-positive persons who have never been on ART.[5] |
| Upper bound | 1.04% | 12% | This is the hypothetical worst-case scenario in which the many persons deteriorate more rapidly. |

Important limitations to note include: (i) we do not model any interaction between HIV and COVID-19 infection – that is, PLHIV are not assumed to be more or less likely to acquire or die from COVID-19; (ii) the effect of disruption on the risk of mother-to-child transmission is not included in this model; (iii) the effects of any social distancing on risk behaviours and new partner acquisition is not included; (iv) possible increases in drug resistance due to ART regimens being disrupted are not included but these could also contribute to excess HIV deaths in the longer term.

## Results

Figure 1 shows the projected number of COVID-19 deaths for the three intervention scenarios from [3], together with the projections for excess HIV-related deaths in South Africa over time for the three service disruption scenarios. The excess HIV-related deaths are spread over the five-year time period, in contrast to the direct COVID-19 deaths, which all occur in 2020-2021.

**Figure 1:**
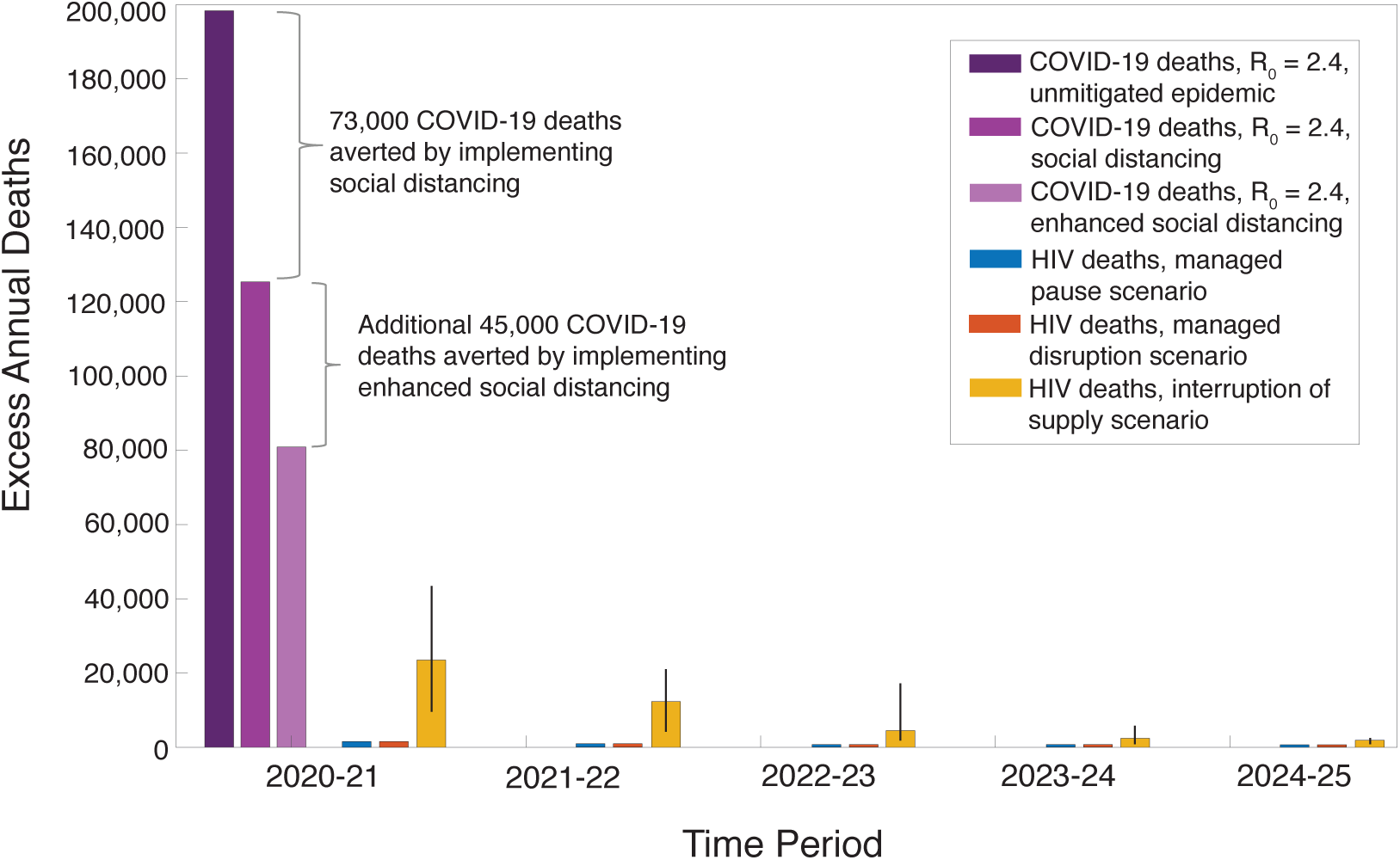
Projected direct and indirect deaths over time, under different assumptions about the COVID-19 epidemic and the impact on HIV services (Table 1), for three months of service interruption. Confidence intervals represent uncertainty in the monthly mortality risk for individuals with interrupted ART, ranging from 0.26%-1.04% per month.

The numbers of additional HIV-related deaths in the less severe scenarios (‘Managed pause’ and ‘Managed disruption’) are small (less than 1%) but could nevertheless lead to a significant increase in new HIV infections (Figure 2), setting back HIV control efforts. For the most severe ‘Interruption of supply’ scenario, 23,000 excess HIV-related deaths in the first year represent a substantial increase on the 71,000 HIV-related deaths estimated in South Africa in 2018.[6] Over five years, the total number of excess deaths caused by the disruptions (38,000) is smaller than those that would be caused by the COVID-19 epidemic itself, but of the same order of magnitude as the reductions in deaths that could be caused by the proposed COVID-19 epidemic mitigation interventions (i.e., social distancing for the whole population (73,000 fewer deaths), or social distancing with enhanced social distancing for the elderly (a further 45,000 fewer deaths)). These excess HIV-related deaths are initially attributed primarily to the increased mortality risk for people stopping ART use, but over five years an increasing proportion are also due to others not having achieved a timely ART initiation during the interruption.

**Figure 2:**
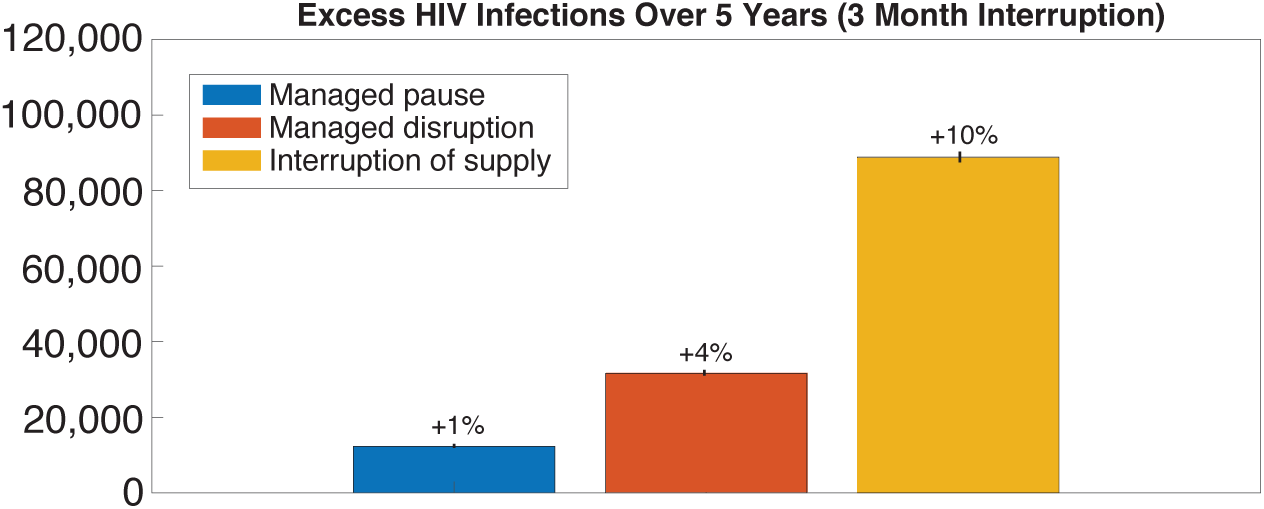
Excess HIV infections in South Africa over 5 years for three scenarios (managed pause = blue bars, managed disruption = orange bars, interruption of supply = yellow bars) for a three month interruption.

We also examined scenarios in which the duration of disruptions and the proportion of PLHIV on ART who are affected were varied and alternative assumptions were made for the mortality risk for persons whose ART is interrupted (Figure 3). Under the lowest mortality assumption, for most scenarios there are fewer additional HIV-related deaths than COVID-19 deaths and fewer HIV-related deaths compared to those that could be saved by the interventions to prevent COVID-19 epidemic spread. However, it is worth noting that under some very severe (and unlikely) circumstances of wide-spread and long interruptions of ART, the number of additional HIV-related deaths could be much greater than our default assumptions suggest.

**Figure 3:**
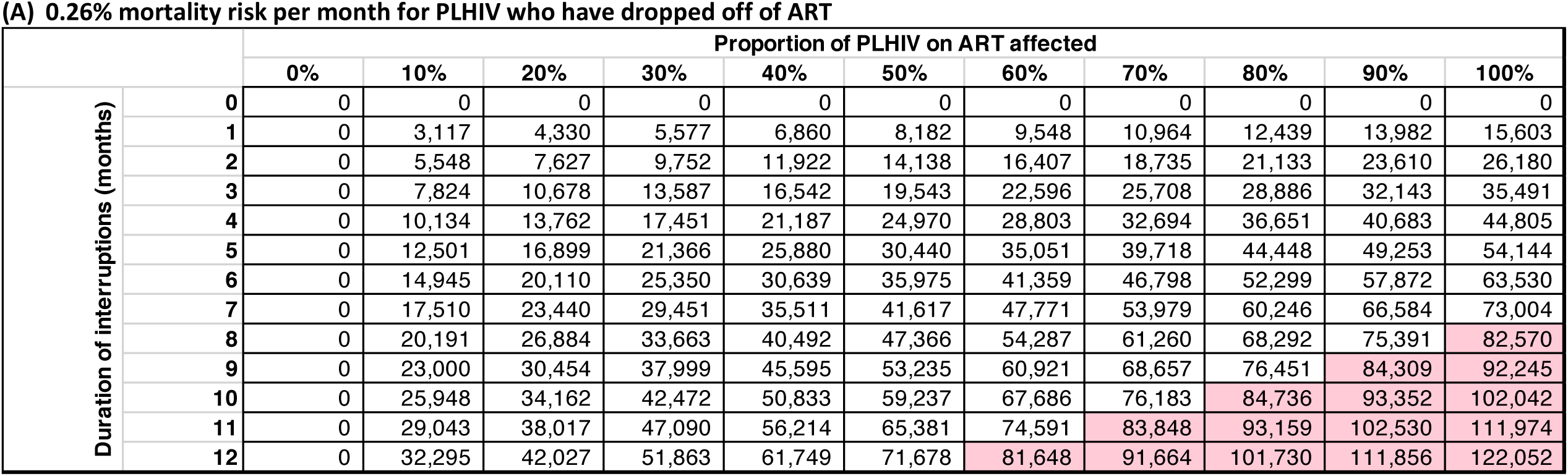
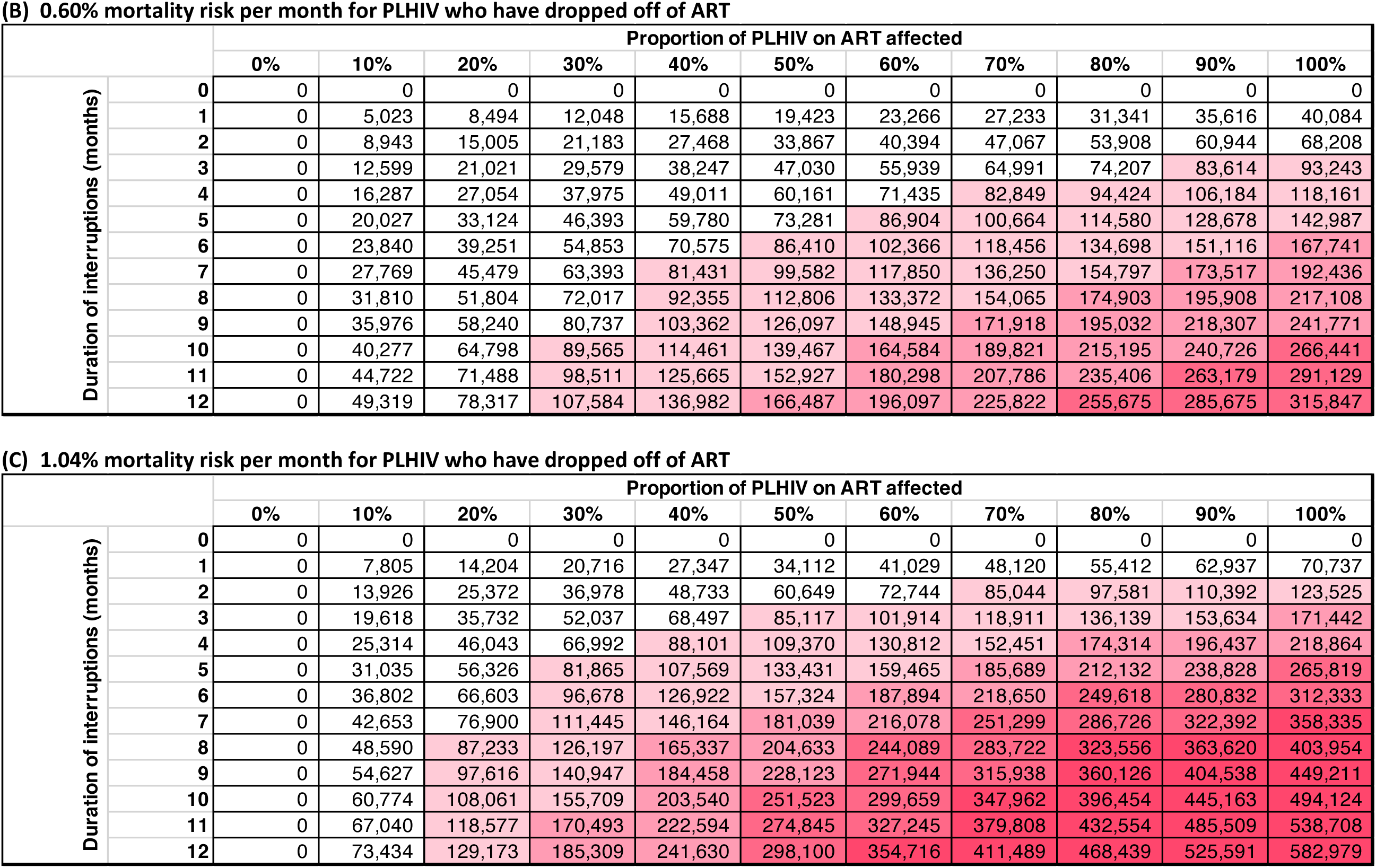
The number of excess cumulative HIV deaths in South Africa over 5 years by the duration of interruption (months) and the proportion of people living with HIV (PLHIV) who are on ART affected by the interruption. Three scenarios are presented for mortality risk per month for PLHIV who have ceased ART due to the interruption: (A) 0.26%, (B) 0.60%, and (C) 1.04%. Colour gradients correspond to deaths greater than the lower number of deaths expected with enhanced social distancing for a COVIDR19 epidemic in Africa with R_0_=2.4. Light pink corresponds to ≥1 times the number of COVIDR19 deaths with the darkest colour corresponding to ≥4 times the number of COVIDR19 deaths.

## Conclusions

Maintaining ART treatment during any health system disruptions during the COVID-19 epidemic is the overriding priority for HIV programs. A wide-spread and long-lasting interruption to ART supply could cause additional deaths on the same order of magnitude as those which could be saved by interventions to mitigate the COVID-19 epidemic itself. Some of the negative impacts of service disruption could be minimised by policy changes that are beneficial to HIV programs in the long-term – for example, adaptations to ART delivery such as multi-month ART scripting are now recommended for consideration in appropriate circumstances by the WHO and PEPFAR.[7, 8] However, if an interruption to ART supplies does occur, it may also take programs longer than assumed here to return to status quo, with further adverse effects expected among individuals who experience long-term ART disruptions.

Some short-term changes to programs, such as scaling back new VMMC and reduced use of condoms, may be less impactful in terms of the excess number of deaths in the medium-term, but we note that new HIV infections would increase and could have further long-term ramifications not represented in the five-year time horizon. The analysis presented here is for South Africa, but it is anticipated that the overall direction of the results will be similar for many countries in sub-Saharan Africa with large HIV epidemics.

The assumptions made are not predictions of the future, but hypothetical scenarios designed to allow this analysis to highlight the importance of maintaining different services during the COVID-19 epidemic. The model itself has many limitations (noted above). The major uncertainties in this analysis can be classified into four groups: (1) uncertainty about the scale of the COVID-19 epidemic (how far and fast will it spread and with what chance of death for those infected); (2) uncertainty into the extent to which HIV programs will actually be disrupted by the COVID-19 epidemic and the response to it; (3) uncertainty about changes to patterns of sexual risk behaviour in response to the epidemic; and (4) uncertainty about the mortality risk of those persons on ART who may suffer an interruption in the supply of drug.

We have not assumed any interaction between HIV or ART status and COVID-19 infection in this analysis. This may need to be revised as more information becomes available, and may influence outcomes particularly in areas with large numbers of PLHIV who are not virally suppressed.

We have also not assumed any changes in sexual risk behaviour. A reduction in sexual contact rates could reduce incidence rates and hence new infections but would have a limited impact on the number of deaths over five years. Furthermore, interruptions to condom supplies could counteract any incidence decline. Reductions in condom availability could also have differential impacts on new HIV infections among different populations; for example, female sex workers might be at higher excess risk than women in stable partnerships. Ensuring support for community prevention programs during the COVID-19 epidemic will help to mitigate these potential effects.

The range of mortality effects due to ART disruption will depend on the health and immune status of the person, the drugs they are using and their treatment history, as well as any steps that are taken to prolong supply (alternate day dosing, for example). For this reason, we have repeated the analysis for a wide range of values, but even within this, it may be that the effect has been over or under-estimated. Ultimately, ensuring the ART supply for individuals currently on treatment would minimise excess mortality among PLHIV in South Africa, and should be a key policy priority.

## Data Availability

There are no data referred to in this study beyond those that are cited.

